# Demand for hospitalization services for COVID-19 patients in Brazil

**DOI:** 10.1101/2020.03.30.20047662

**Authors:** Marcia C. Castro, Lucas Resende de Carvalho, Taylor Chin, Rebecca Kahn, Giovanny V. A. França, Eduardo Marques Macário, Wanderson Kleber de Oliveira

## Abstract

COVID-19 is now a pandemic and many of the affected countries face severe shortages of hospital resources. In Brazil, the first case was reported on February 26. As the number of cases grows in the country, there is a concern that the health system may become overwhelmed, resulting in shortages of hospital beds, intensive care unit beds, and mechanical ventilators. The timing of shortage is likely to vary geographically depending on the observed onset and pace of transmission observed, on the availability of resources, and on the actions implemented. Here we consider the daily number of cases reported in municipalities in Brazil to simulate twelve alternative scenarios of the likely timing of shortage, based on parameters consistently reported for China and Italy, on rates of hospital occupancy for other health conditions observed in Brazil in 2019, and on assumptions of allocation of patients in public and private facilities. Results show that hospital services could start to experience shortages of hospital beds, ICU beds, and ventilators in early April, the most critical situation observed for ICU beds. Increasing the allocation of beds for COVID-19 (in lieu of other conditions) or temporarily placing all resources under the administration of the state delays the anticipated start of shortages by a week. This suggests that solutions adopted by the Brazilian government must aim at expanding the available capacity (e.g., makeshift hospitals), and not simply prioritizing the allocation of available resources to COVID-19.

## Introduction

On March 11 the World Health Organization characterized COVID-19 as a pandemic. Caused by the novel coronavirus SARS-CoV-2, it emerged in China and quickly spread across the country and beyond. As of March 27, it was present in 202 countries and territories, with 509,164 cases and 23,335 deaths reported.^1^ The clinical course of COVID-19 poses serious challenges to the health system and may call for drastic rationing decisions.^2,3^ Specifically, allocating hospital beds, intensive care unit (ICU) beds, and mechanical ventilators to COVID-19 patients, besides the ongoing demand driven by other conditions, is a real or soon to be a real problem.^4-6^

Brazil recorded the first COVID-19 case on February 26 and the first death on March 17, both in São Paulo. In 24 days, the disease had spread to every federal unit. As of March 27, 3,417 cases and 92 deaths had been reported. As the numbers increase across the country, so does the demand for hospital services, raising concerns of hospital capacity to cope with that demand. Brazil has a hybrid health system.^7,8^ On the one hand, every citizen has free access to the Unified Health System (SUS). On the other hand, the private sector offers services covered by out of pocket payments and private insurance plans. About 80% of the Brazilian population relies solely on SUS, but this number varies widely across the country reflecting striking inequalities.^9^ The offer of hospital services is also unequal. As of December 2019, 67% and 48% of the total available hospital beds and ICU beds, respectively, were offered in the public system.Geographically, the distribution was also unequal, with 9 and 21 beds per 100,000 people in the North and Southeast regions, respectively.^10^

Here we address this concern and simulate the time it would take for hospitals to operate at capacity, given the current trajectory of COVID-19 in Brazil. We considered different scenarios of supply and demand and detailed the simulations by day and health macro-region. Since not all services are available in each municipality, the Ministry of Health (MoH) considers the macro-region to guide the regionalization of health services.

## Methods

### DATA SOURCES

Population by age group and the number of confirmed COVID-19 cases were obtained from the MoH.^11^ Information on available hospital beds, ICU beds, and mechanical ventilators as of December of 2019 was extracted from the National Registry of Health Establishments (CNES, http://cnes.datasus.gov.br/). We excluded pediatric and obstetric beds, and pediatric, neonatal, and burn recovery ICU beds. We distinguished hospital beds and ICU beds by type of service (private and public); this classification was not available for mechanical ventilators. Average public hospital occupancy in 2019 (detailed by hospital beds and ICU beds) was obtained from the hospitalization system of the MoH (http://sihd.datasus.gov.br/). There was no information on the occupancy rate for private hospitals (here we assumed the same rate obtained for public hospitals). Lastly, access to private health insurance in December 2019 was extracted from the Brazilian Regulatory Agency (http://www.ans.gov.br/perfil-do-setor/dados-e-indicadores-do-setor).

### STATISTICAL ANALYSIS

We conducted forward simulations of the demand for hospital beds, ICU beds, and mechanical ventilators by health macro-region in Brazil. In the initial phase of a new infectious disease outbreak, the number of cases grows exponentially.^12,13^ Using the number of cases reported to date and estimates of doubling times from the outbreak in China,^14-16^ we estimated the number of future cases each day. As the outbreak spreads, the exponential model becomes less realistic,^13^ so we stopped the simulations once 10% of the population was estimated to be infected. We used the age structure of each macro-region combined with age-specific attack and severity rates drawn from the literature. We used attack rates by age from the outbreak in China,^17^ considering a range of ±3%, and considered that 86% of all infections were undocumented.^18^ Demand for hospitalization was calculated using the severity of cases by age from China.^17^ Time from illness to hospitalization was 3-7 days,^19,20^ and length of hospitalization ranged from 7-15 days;^21^ for ICU hospitalizations these parameters were 8-15,^19,20^ and 7-15,^21,22^ respectively. We considered that 5% of the cases needed ICU admission^21,23^ and half of those in ICU needed mechanical ventilation^20,21^ for an average of 5 days.^24^ We also simulated an alternative scenario based on Italy and assumed that 12% of the cases needed ICU admission.^4,6^ These parameters were combined with assumptions of hospital occupancy and type of service to produce twelve scenarios, summarized in Table 1. We performed 1,000 runs for each scenario, drawing from the ±3% uncertainty around attack rates, and considered the results that represented the median of the distribution. All analyses were performed with the use of R software, version 3·6·3, and Rstudio, version 1·2·5033 (R Foundation for Statistical Computing).

**Table 1.**
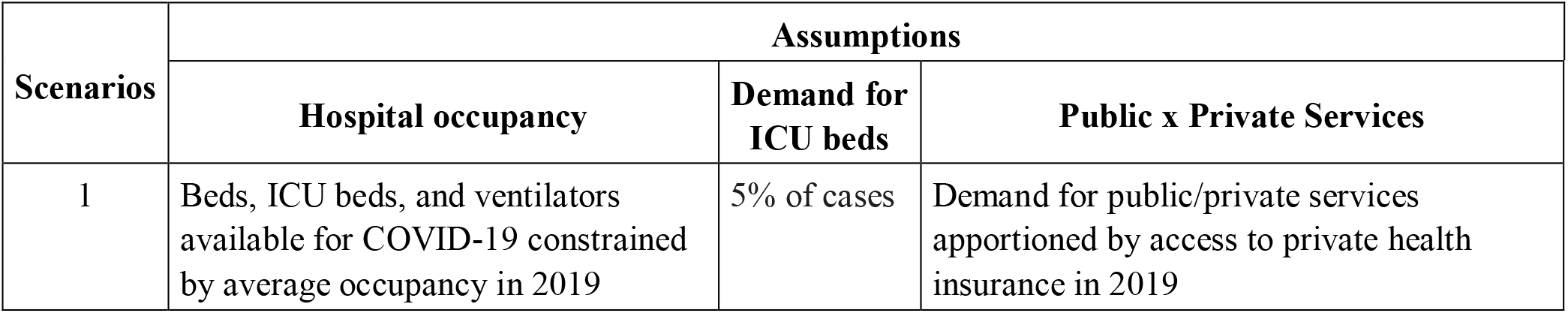
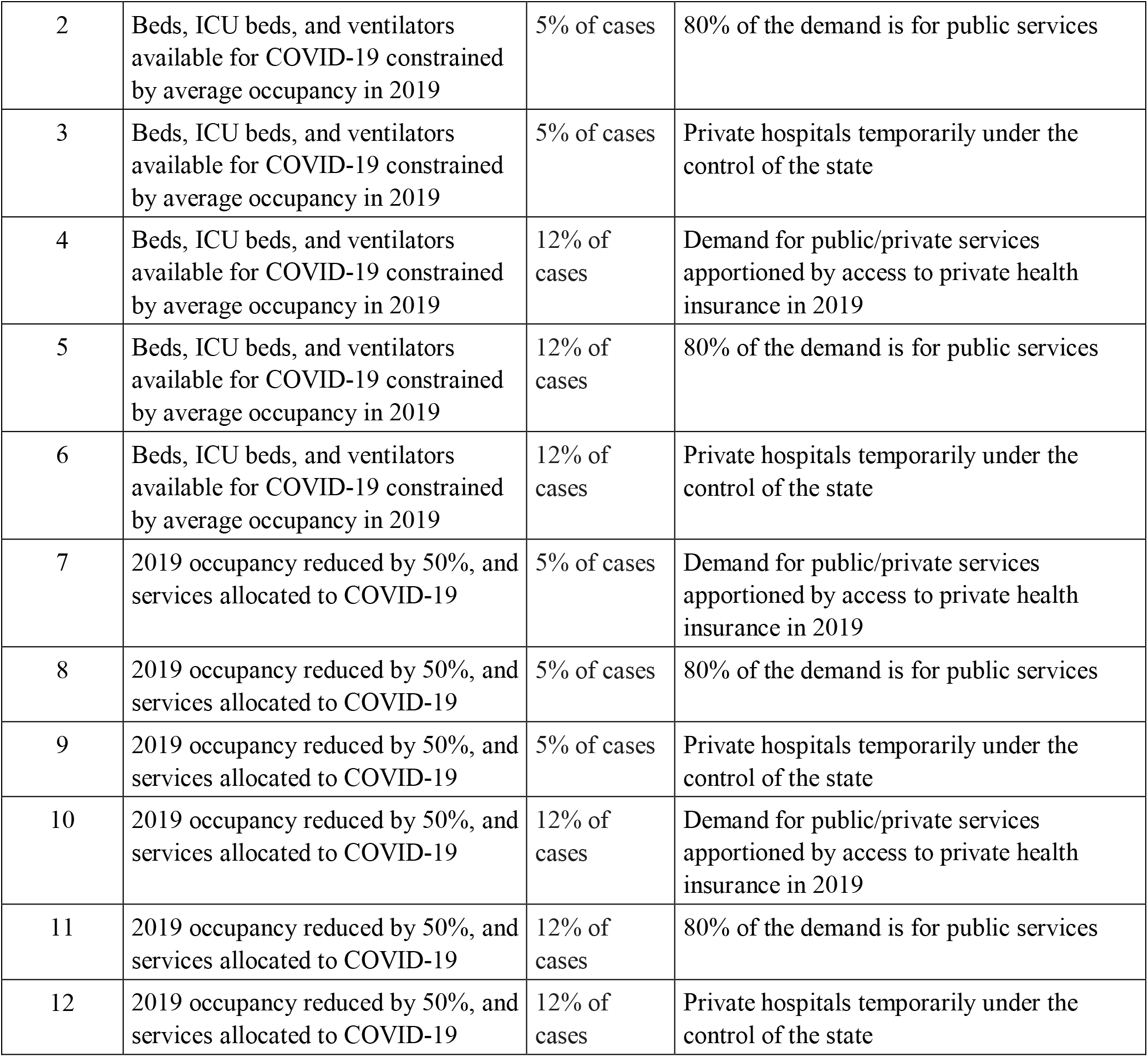
Parameters considered for simulated scenarios

| Scenarios | Assumptions |  |  |
| --- | --- | --- | --- |
|  | Hospital occupancy | Demand for ICU beds | Public x Private Services |
| 1 | Beds, ICU beds, and ventilators available for COVID-19 constrained by average occupancy in 2019 | 5% of cases | Demand for public/private services apportioned by access to private health insurance in 2019 |
| 2 | Beds, ICU beds, and ventilators available for COVID-19 constrained by average occupancy in 2019 | 5% of cases | 80% of the demand is for public services |
| 3 | Beds, ICU beds, and ventilators available for COVID-19 constrained by average occupancy in 2019 | 5% of cases | Private hospitals temporarily under the control of the state |
| 4 | Beds, ICU beds, and ventilators available for COVID-19 constrained by average occupancy in 2019 | 12% of cases | Demand for public/private services apportioned by access to private health insurance in 2019 |
| 5 | Beds, ICU beds, and ventilators available for COVID-19 constrained by average occupancy in 2019 | 12% of cases | 80% of the demand is for public services |
| 6 | Beds, ICU beds, and ventilators available for COVID-19 constrained by average occupancy in 2019 | 12% of cases | Private hospitals temporarily under the control of the state |
| 7 | 2019 occupancy reduced by 50%, and services allocated to COVID-19 | 5% of cases | Demand for public/private services apportioned by access to private health insurance in 2019 |
| 8 | 2019 occupancy reduced by 50%, and services allocated to COVID-19 | 5% of cases | 80% of the demand is for public services |
| 9 | 2019 occupancy reduced by 50%, and services allocated to COVID-19 | 5% of cases | Private hospitals temporarily under the control of the state |
| 10 | 2019 occupancy reduced by 50%, and services allocated to COVID-19 | 12% of cases | Demand for public/private services apportioned by access to private health insurance in 2019 |
| 11 | 2019 occupancy reduced by 50%, and services allocated to COVID-19 | 12% of cases | 80% of the demand is for public services |
| 12 | 2019 occupancy reduced by 50%, and services allocated to COVID-19 | 12% of cases | Private hospitals temporarily under the control of the state |

## Results

Figure 1 shows the fraction of public hospital beds and ICU beds in each health micro-region, as well as the percentage of the population in those units that rely solely on the SUS. Inequities in supply and demand exist, and mirror regional inequalities commonly found in social and health indicators.^9^

**Figure 1.**
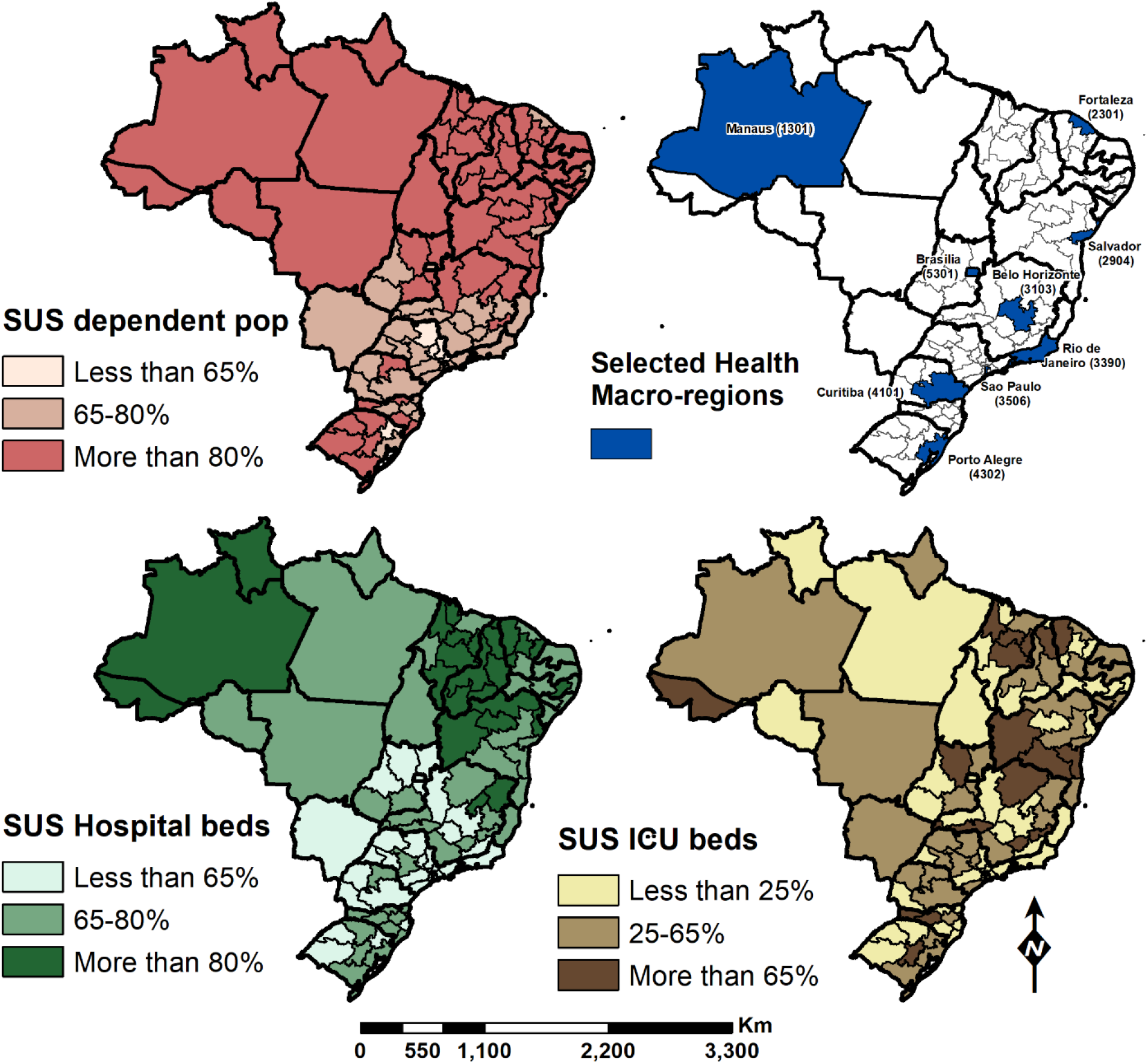
Percentage of the population that relies solely on the SUS, percentage of beds available in each health macro-region that are allocated to, and location of the simulated micro-regions. Thicker black lines define the boundaries of the federal units. Selected health macro-regions are named after the capital city, and the ID code is also provided.

As of March 27, 3.417 cases were confirmed in Brazil, 70% concentrated in nine cities: São Paulo, Rio de Janeiro, Fortaleza, Brasília, Porto Alegre, Salvador, Belo Horizonte, Curitiba, and Manaus. The growth pattern of the epidemic is not the same across the country. Therefore, we chose to simulate scenarios for the nine health macro-regions associated with those cities; combined these macro-regions represent 79·6% of reported cases. In the case of São Paulo and Brasília, the macro-region includes only the municipality itself. Rio de Janeiro and Manaus have only one macro-region for the entire state. Fortaleza, Porto Alegre, Belo Horizonte, Salvador, and Curitiba are in macro-regions that comprise 44, 89, 103, 48, and 93 municipalities, respectively.

Our simulated scenarios indicate that hospital services could start to experience shortages of hospital beds, ICU beds, and ventilators across macro-regions as early as April (Figure 2). ICU beds are, by far, the most pressing need. The supply of hospital beds is likely to face shortages after April 21 and of mechanical ventilators just a few days afterward. The two scenarios for ICU admission (5% and 12% of the total cases) resulted in about 4-6 days difference. With a 12% assumption, Brasília would face a shortage of ICU beds by the end of March. Making 50% of usual occupancy dedicated to COVID-19 postpones shortages for less than a week, and placing all resources under state control shifts the capacity threshold by up to one week for hospital beds and mechanical ventilators, and by up to four days for ICU beds. Considering alternative hypothesis of access to public resources, scenarios 2, 5, 8, and 11 assumed that 80% of the demand would come from individuals that solely rely on SUS. Since the offer of SUS services presents regional inequities (Figure 1), the timing of shortage depends both on the share public x private in each area.

**Figure 2.**
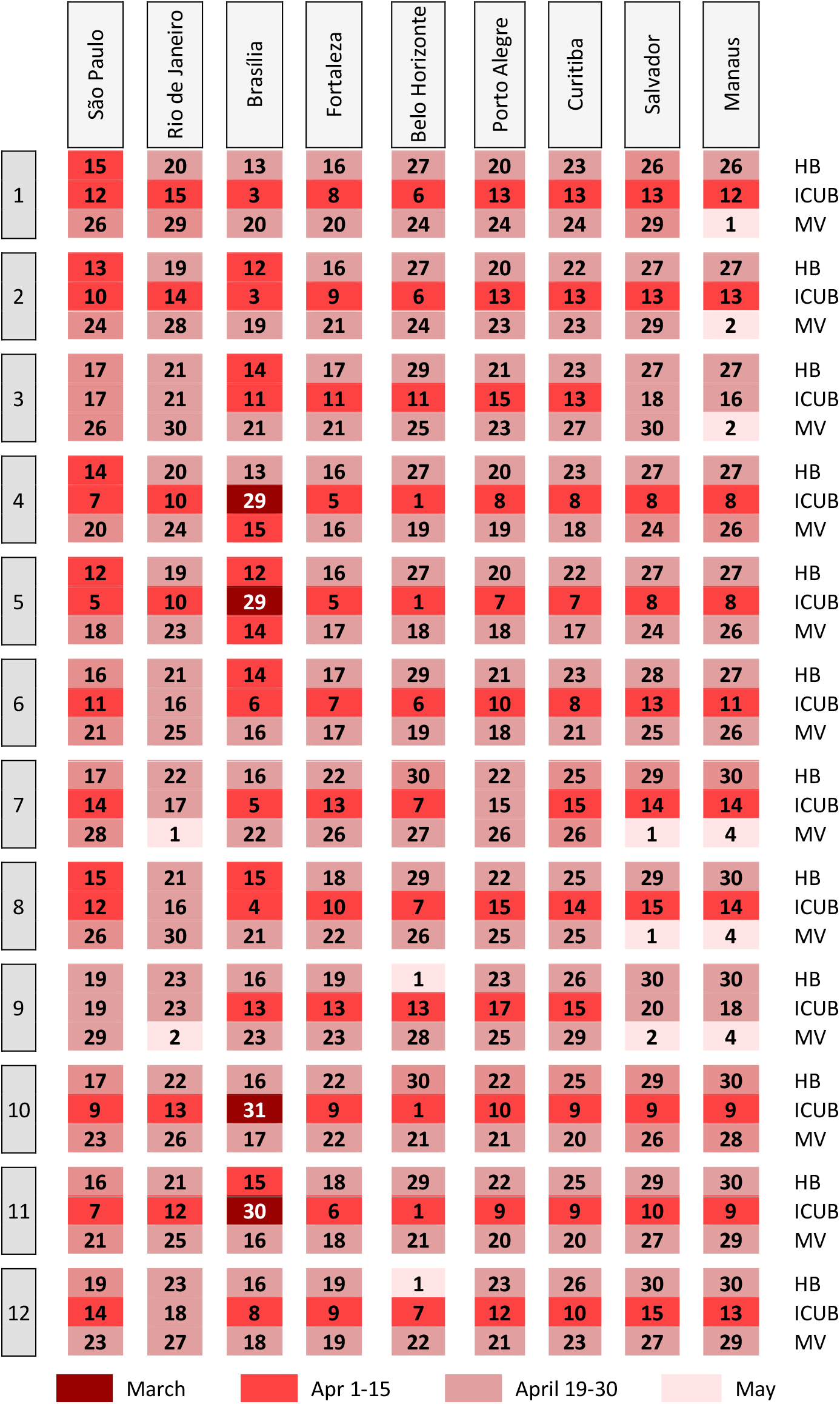
Date when hospital services will be at capacity in each simulated scenario. Numbers on the left represent each of the 12 simulated scenarios described in Table1. Names on top indicate the municipality of reference to the health macro-region. HB = hospital beds. ICUB = ICU beds. MV = mechanical ventilators. Numbers on the table refer to day of the month, and cell colors indicate the month. Results reflect the median after 1,000 runs of the model.

The timing of shortage varied by health macro-region. It depends on the observed onset and pace of transmission. For example, although the first case in Fortaleza was reported on March 15, 18 days after the first case was reported in the country, it took eight days to reach to 100 cases. In São Paulo, where the first case was reported, the first 100 cases were recorded in 19 days.

## Discussion

Our simulated scenarios show that, if no change in trajectory is observed, around early April Brazil would start to face shortages of hospital beds, ICU beds, and ventilators, with ICU beds being the most immediate problem. The timing of shortages across the country depends on the onset and the intensity of transmission. Also, the population that relies solely on SUS may bear the largest burden, further exacerbating the existing inequalities, which calls for a reflection around equity and ethics in service allocation. Avoiding this scenario is the paramount task of the MoH.

It is unreasonable to expect that all hospital resources could be entirely dedicated to COVID-19. Although elective procedures can be postponed, other health emergencies compete for resources. For example, dengue transmission is presently intense and increasing; until March 14 there were 390,684 cases and 106 deaths (60·4% of them among individuals aged 60 or more). In 2019, with more than 1·5 million cases of dengue reported, 4% (n=55,235) required hospitalization. Most importantly, 45% of the dengue hospitalizations occurred between March and May (21% just in May, the month when most likely the hospitals will be operating at capacity because of COVID-19). In addition, the influenza season is starting, and in 2019 the peak of hospitalizations occurred in May. Besides beds and ventilators, the stress imposed on the health system compromises the overall care, which may result in deterioration of the treatment of other conditions followed by increases in incidence. This scenario was observed during the 2013-14 Ebola outbreak.^25^

Shifts in the current trajectory of the epidemic in Brazil may anticipate or postpone the stress on health services. Importantly, our results are sensitive to assumptions about key parameters in the model, including the doubling time, underreporting rate, age-specific attack and severity rates, time from illness to hospitalization and ICU, and length of stay estimates. By drawing on available evidence of how COVID-19 has and is evolving across the globe, our results rely on and reflect the limitations of the parameters observed in China and on unique patterns reported in Italy. Given the significant uncertainty in parameter estimates at this phase of the epidemic and the simplifying assumptions of an exponential model, results from our simulated scenarios should not be used as precise estimates for the exact timing and extent of capacity thresholds being reached. Instead, they aim to inform planning and prompt response and do send three important messages.

First, the epidemic of COVID-19 is likely to exacerbate existing inequalities if those that solely rely on the SUS are hit the hardest. In communities with high population density and poor infrastructure, it is unfeasible to practice social isolation. In those areas, the transmission is likely to occur fast and the population mostly relies only on the SUS, quickly overwhelming the health system. One alternative to alleviate the is to temporarily put all private hospitals under the control of the state, a measure adopted by Spain.^26^ Yet, our results show that this would postpone shortages by about a week. In fact, solutions that do not involve the opening of new facilities will have a very short effect on the timing of shortage.

Second, there is a short window of opportunity to prepare. The response must be immediate, and it will demand a concerted effort from society. Repurposing large spaces (e.g., arenas, convention centers) to build makeshift hospitals for additional beds is critical. Calling on the industry to produce the necessary equipment (e.g., ventilators, masks, gloves, protective gown, etc) and to provide it to hospitals at minimum or no cost would also contribute to mitigating the stress on the system, and to safeguarding the working conditions of health care professionals.

Mobilizing community leaders, artists, athletes, and local role models would help to convey a unique message to the population, and to gather support from the wealthy that could provide resources to expand the production of medical equipment.

Third, additional actions to contain the transmission and to change the current trajectory of the epidemic must be taken. Partial shutdown has been initiated in many Brazilian cities, but compliance is far from ideal. Increasing the availability of tests would allow the government to comprehensively trace and test all contacts of positive cases, a strategy successfully adopted by South Korea.^27^ Without intensified actions, or even worse, if current actions are loosened, Brazil may face the need to implement guidelines for rationing health resources,^2^ which invariantly call for difficult decisions.

## Conclusion

Brazil is, in theory, uniquely equipped to respond to the COVID-19 epidemic. It has a free and universal health system,^7^ it has one of largest community-based primary care delivery programs that serves 74·8% of the population,^28^ it can learn from the mistakes and success that other countries hit by COVID-19 have made, and it has a history of responding to health threats by implementing governmental action and by generating high-quality scientific evidence, such as was done when Zika virus hit the country.^29^ Yet, the current moment is unique. It requires a unified message from the country’s leadership at various levels: federal, state, and municipal. It requires the industry to work in solidarity to produce needed inputs without aiming profit but the collective wellbeing. It requires the population to realize the importance and the urgency to comply. We hope our results will help to move forward this agenda.

## Data Availability

Code is available on github.

https://github.com/lucasrdc/covid19/blob/master/covid19.R

## Contributors

MCC conceived the original, was responsible for data analysis, data interpretation, data visualization, and wrote the manuscript. LRC was responsible for data curation, data analysis, programming, and contributed to writing. RK and TC were responsible for programming and contributed to writing. GVAF, EMM, and WKO were responsible for data curation and interpretation. All authors approved the final version of the manuscript.

